# Ejection fraction quantification from ungated chest CT by AI

**DOI:** 10.1101/2025.11.14.25339933

**Authors:** Jianhang Zhou, Jacek Kwieciński, Aakash Shanbhag, Konrad Pieszko, Giselle Ramirez, Mark Lemley, Aditya Killekar, Waseem Hijazi, Robert J.H. Miller, Paul B. Kavanagh, Joanna X. Liang, Leandro Slipczuk, Mark I. Travin, Erick Alexanderson, Isabel Carvajal-Juarez, René R.S. Packard, Mouaz Al-Mallah, Andrew J. Einstein, Wanda Acampa, Stacey Knight, Viet T Le, Steve Mason, Thomas L. Rosamond, Jarosław Hiczkiewicz, Samuel Wopperer, Panithaya Chareonthaitawee, Daniel S. Berman, David. E. Newby, Marcelo F. Di Carli, Damini Dey, Piotr J Slomka

## Abstract

Left ventricular ejection fraction (LVEF) is an important clinical metric, obtained by specialized imaging across the cardiac cycle. We present a novel AI approach to estimate LVEF from ungated chest CT. Using multicenter (11 sites) registry of 25,852 patients, AI-derived CT LVEF (AI LVEF) showed strong correlation with 3D gated positron emission tomography (r=0.84), area under the curve (AUC) of 0.96, negative predictive value of 95% for reduced LVEF (< 40%), and effectively stratified risk of heart failure, cardiovascular death, and all-cause death. In a separate large multicenter population (n=24,054) with lung CT scans, reduced AI LVEF was associated with a hazard ratio of 13.3 (95% confidence interval 9.7-18.4) for cardiovascular death. AI LVEF could also predict reduced echocardiographic LVEF (AUC=0.91).

LVEF can be accurately estimated from non-contrast, ungated, low-dose chest CT scans, effectively stratifying patients for heart failure and mortality, with potential widespread clinical utility.

---

Left ventricular ejection fraction (LVEF) remains a cornerstone metric of cardiac function for diagnosing heart failure, stratifying risk, and selecting guideline-directed medical and device therapies.^1,2^ It defines heart failure (HF) subtypes: preserved (≥ 50%), mildly reduced (40-49%),^3^ or reduced (< 40%) LVEF, which guides management and prognostication.^4^ LVEF measurements underpin thresholds for therapeutic interventions, from pharmacotherapy and device implantation to disability assessment or perioperative planning.

Computed tomography (CT) is the most widely used cross-sectional imaging modality in global clinical practice. Chest CT scans are performed ubiquitously for lung disease evaluation, cancer screening, trauma, pulmonary embolism, and other indications. Over 20 million chest CTs were performed in the US in 2023.^5^ Accurate evaluation of LVEF from these scans would enhance cardiovascular screening, enabling early identification of systolic dysfunction in populations undergoing chest CT for non-cardiac indications (e.g., smokers, oncology surveillance, trauma), without incurring additional costs, radiation, or contrast exposure. Early detection of subclinical dysfunction could prompt timely echocardiographic confirmation, optimize therapy, and ultimately improve prognosis. However, few studies have addressed the possibility of estimating quantitative LVEF values from non-contrast, ungated chest CT.

Here, we present a novel machine-learning framework that combines deep learning and a boosting regressor to estimate continuous LVEF values from static chest CT. Using a unique multicenter registry of cardiac PET/CT data that includes ultra-low-dose ungated chest CT performed for attenuation correction, we validate our CT-based prediction against LVEF derived from PET scans and evaluate its performance across clinically meaningful LVEF thresholds. We further validate the AI LVEF with echocardiographic LVEF and demonstrate its prognostic value in the National Lung Screening Trial (NLST). The hybrid approach is interpretable at the individual patient level, informing physicians which features are most important for estimating LVEF. Our approach demonstrates, for the first time, that quantitative LVEF can be derived from ungated chest CT scans.

## Results

### Overview of the study

The workflow of AI LVEF prediction from non-contrast ungated chest CT is illustrated in **Figure 1**. From 11 international imaging centers of cohort 1, we included 25,852 patients with hybrid PET/CT scans. The reference LVEF was derived from gated cardiac PET scans. We used previously established deep-learning frameworks^6–8^ to segment CT structures: left ventricular myocardium, left and right ventricles, left and right atria, left and right lungs, aorta, pulmonary artery, coronary artery calcium (CAC), and epicardial adipose tissue (EAT). Machine-learning regression models were developed to integrate measurements of these structures, along with heart rate, blood pressure, age, sex, height, and weight. A total of 11 models, each built from 10 sites, were externally tested using the leave-one-site-out strategy in the 11^th^ site. We evaluated the AI LVEF prediction both as a continuous variable and after applying clinical thresholds, as well as in the survival analysis of HF and death. We also applied the model developed using the entire cohort 1 to cohorts 2 and 3. In cohort 2, we compared AI LVEF with LVEF derived from echocardiography and assessed agreement. In cohort 3, AI LVEF was used to stratify the risks of cardiovascular and all-cause death in subjects from the NLST.

**Figure 1.**
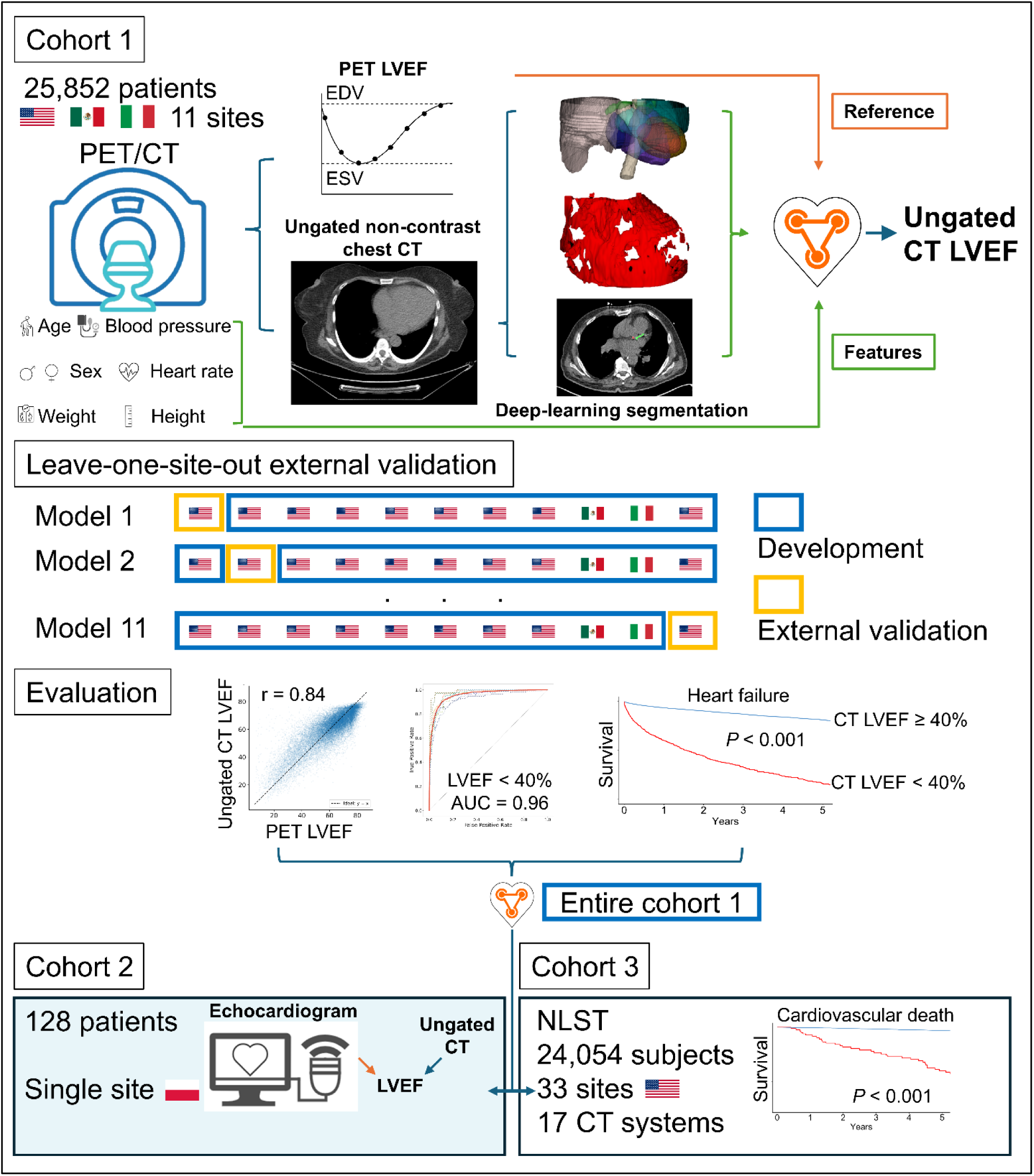
AI LVEF workflow Overview: Cohort 1 included a total of 25,852 patients from 11 geographically separate medical centers (3 countries). Left ventricular ejection fraction (LVEF) was derived from end-diastolic volume (EDV) and end-systolic volume (ESV) in gated cardiac PET and used as the reference for CT-wise artificial intelligence (AI)-enabled estimation that integrated deep-learning CT structures and basic clinical variables. We conducted leave-one-site-out external validation and created 11 models. For each model, one site was reserved for external validation, and the remaining 10 sites were used for model training. This process was repeated 11 times across the entire registry-resulting in 11 independent models and 11 independent external evaluations. We evaluated AI LVEF prediction using both continuous and binary metrics, as well as survival analysis. The model developed using cohort 1 was applied to cohort 2 for comparison with echocardiographic LVEF (n = 128) and to cohort 3 for risk stratification in the National Lung Screening Trial (NLST, n = 24,054).

### Baseline characteristics

The baseline characteristics of cohort 1 are shown in **Table 1**. The mean age was 66 ± 13 years, and 41% were female. The median resting heart rate was 69 beats per minute (bpm) (interquartile interval [IQI] 61 bpm, 78 bpm), with median resting systolic and diastolic pressures of 132 mmHg (IQI 119 mmHg, 148 mmHg) and 70 mmHg (IQI 61 mmHg, 79 mmHg), respectively. The median LVEF was 64% (IQI 54%, 72%), with 11% having LVEF < 40%.

**Table 1.**
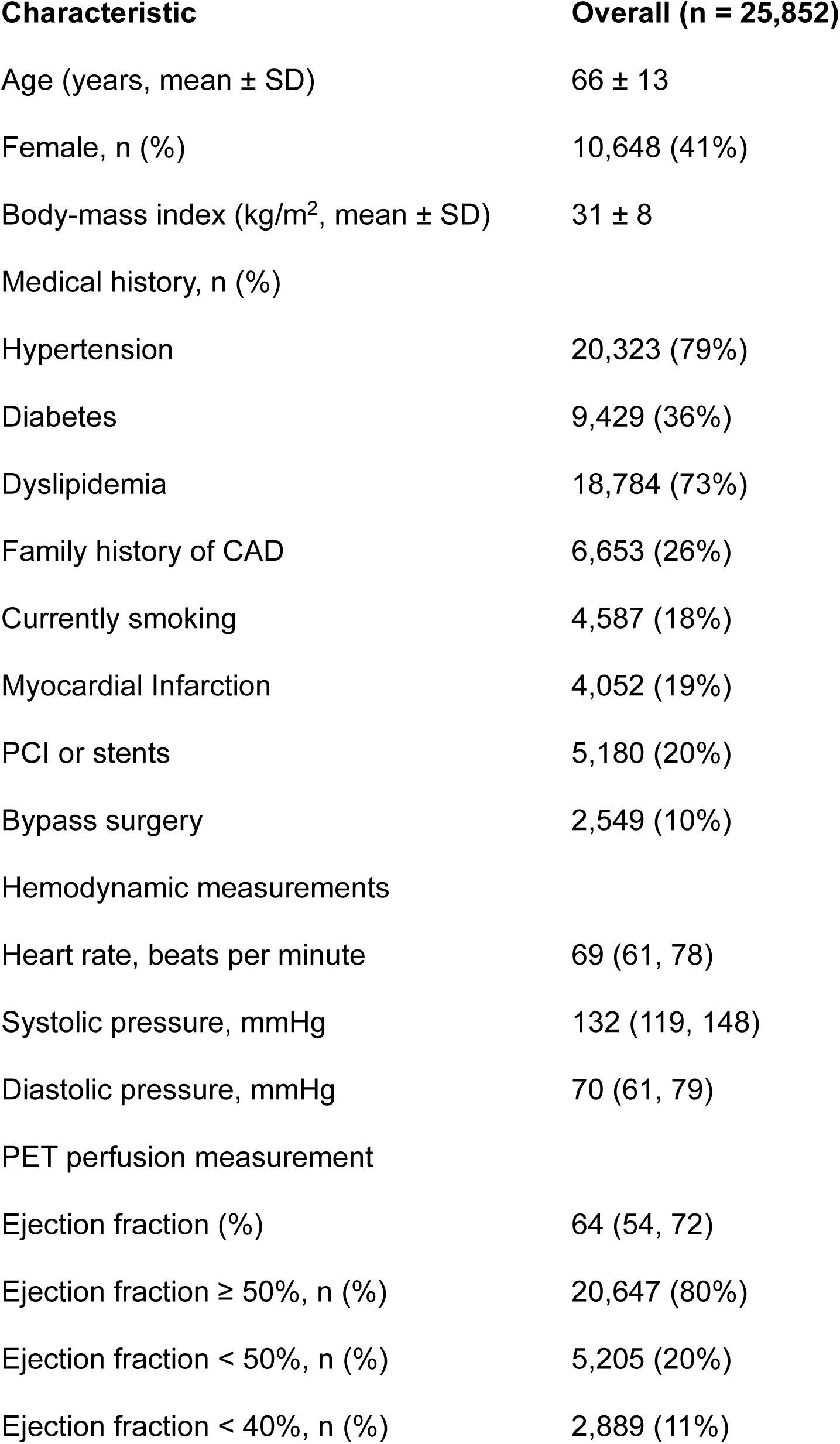
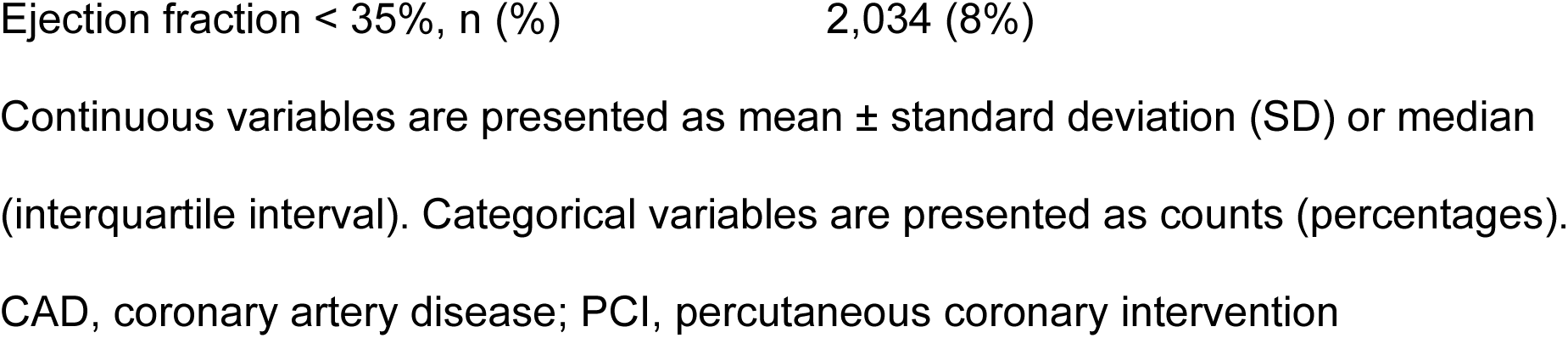
Baseline characteristics of cohort 1.

| **Characteristic** | **Overall (n = 25,852)** |
| --- | --- |
| Age (years, mean ± SD) | 66 ± 13 |
| Female, n (%) | 10,648 (41%) |
| Body-mass index (kg/m<sup>2</sup>, mean ± SD) | 31 ± 8 |
| Medical history, n (%) |  |
| Hypertension | 20,323 (79%) |
| Diabetes | 9,429 (36%) |
| Dyslipidemia | 18,784 (73%) |
| Family history of CAD | 6,653 (26%) |
| Currently smoking | 4,587 (18%) |
| Myocardial Infarction | 4,052 (19%) |
| PCI or stents | 5,180 (20%) |
| Bypass surgery | 2,549 (10%) |
| Hemodynamic measurements |  |
| Heart rate, beats per minute | 69 (61, 78) |
| Systolic pressure, mmHg | 132 (119, 148) |
| Diastolic pressure, mmHg | 70 (61, 79) |
| PET perfusion measurement |  |
| Ejection fraction (%) | 64 (54, 72) |
| Ejection fraction ≥ 50%, n (%) | 20,647 (80%) |
| Ejection fraction < 50%, n (%) | 5,205 (20%) |
| Ejection fraction < 40%, n (%) | 2,889 (11%) |

Continuous variables are presented as mean ± standard deviation (SD) or median (interquartile interval). Categorical variables are presented as counts (percentages).
CAD, coronary artery disease; PCI, percutaneous coronary intervention

The baseline characteristics of cohort 2 are shown in **Supplementary Table 1**. The mean age was 67 ± 10 years, and 44% were female. The median resting heart rate was 74 bpm (IQI 64 bpm, 78 bpm), with median resting systolic and diastolic pressures of 130 mmHg (IQI 120 mmHg, 140 mmHg) and 80 mmHg (IQI 70 mmHg, 85 mmHg), respectively. The median resting ejection fraction was 55% (IQI 50%, 60%), with 14% having LVEF < 40%.

### Ejection fraction estimation

The performance of cohort 1 in continuous LVEF estimation is shown in **Figure 2**. The mean bias was 0.1% (95% confidence interval [CI]: 0.01%-0.2%), with a mean absolute error (MAE) of 6.0% (95% CI: 5.9%-6.0%) in absolute LVEF units (**Figure 2, a & b**). The Pearson r was 0.844 (95% CI: 0.841-0.848), the intraclass correlation coefficient (ICC) was 0.832 (95% CI: 0.828-0.836), and the coefficient of variation was 13.0 (95% CI: 12.9-13.2), indicating a good agreement with the reference values (**Figure 2, c-e**). Site-level analyses yielded consistent results, with bias ranging from 0 to 0.4%, MAE from 5.1% to 6.2%, r from 0.77 to 0.88, ICC from 0.75 to 0.88, and coefficient of variation from 11.0 to 14.1 across individual sites, supporting the stability and reliability of quantification across imaging centers.

**Figure 2.**
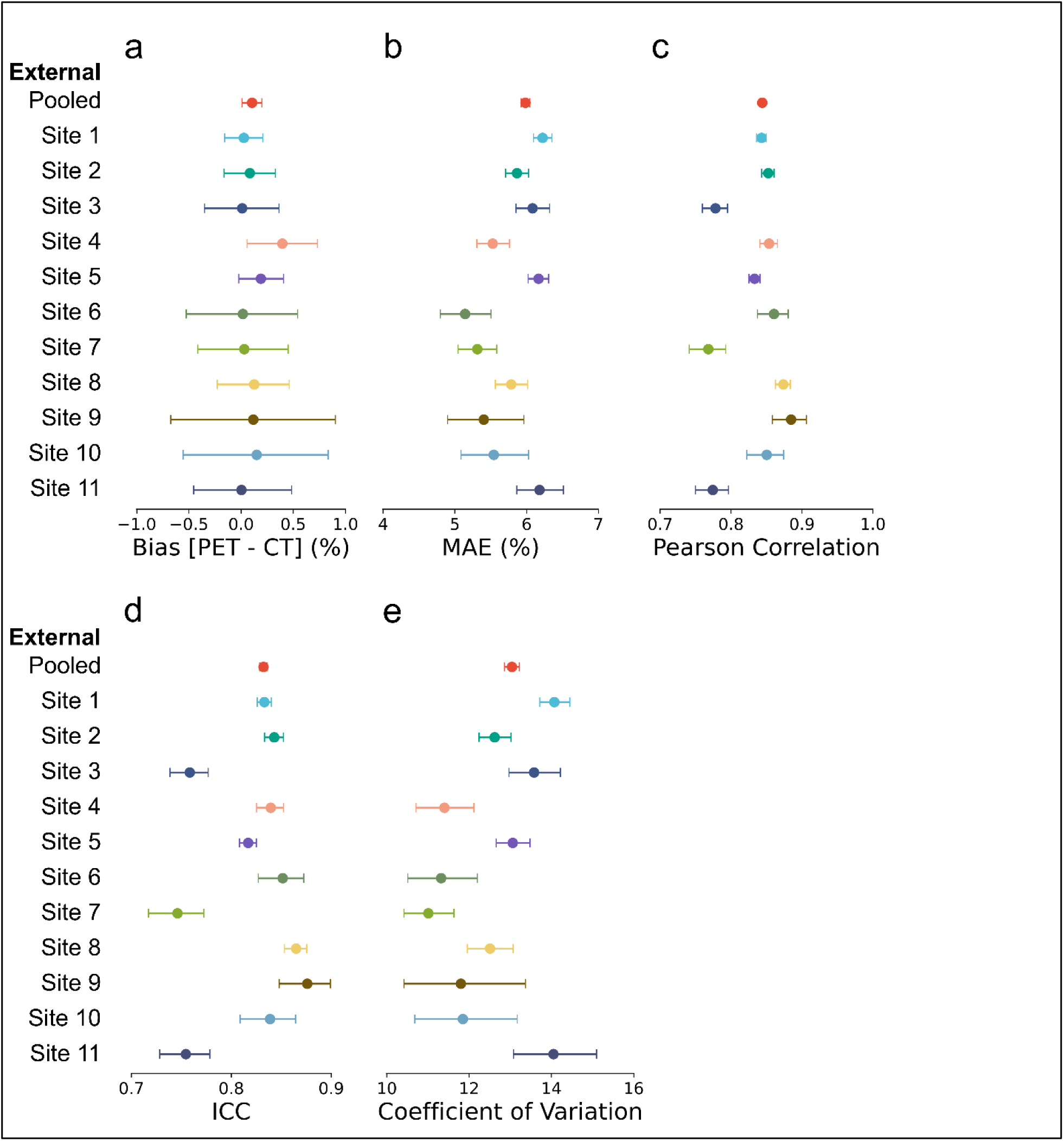
Performance metrics for continuous ejection fraction prediction Continuous left ventricular ejection fraction (LVEF) derived from ungated chest CT was compared with PET LVEF by assessing (a) the bias, (b) the mean absolute error (MAE), (c) the Pearson correlation coefficient, (d) the intraclass correlation coefficient (ICC), and (e) the coefficient of variation in the pooled and site-level external validation with 95% confidence intervals.

In cohort 2, the mean bias between echocardiographic and AI LVEF was -4.2% (95% CI: -5.8% - -2.7%), with a mean absolute error (MAE) of 8.0% (95% CI: 7.0%-9.1%) in absolute LVEF units. The Pearson r was 0.719 (95% CI: 0.623-0.793), the intraclass correlation coefficient (ICC) was 0.657 (95% CI: 0.546-0.745), and the coefficient of variation was 16.8 (95% CI: 14.6-19.1), indicating a moderate agreement with the echocardiographic LVEF. No comparison of LVEF values was tested in cohort 3 due to a lack of ground truth.

### Prediction of reduced CT ejection fraction

Of 25,852 patients in cohort 1, 5,205 (20%) had PET LVEF below 50%, 2,889 (11%) had PET LVEF below 40%, and 2,034 (8%) had PET LVEF below 35% (**Supplementary Table 2**). Site-specific distributions showed variability in the prevalence of reduced LVEF. The proportion of patients with PET LVEF < 50% ranged from 12% to 25%, while the proportion of patients with PET LVEF < 40% ranged from 4% to 15%. Similarly, the PET LVEF < 35% ranged from 1% to 11%.

The discriminative performance of AI LVEF < 40% was excellent, with a pooled AUC of 0.957 (95% CI: 0.953–0.960) (**Figure 3, a & b)**. The overall positive and negative predictive values (PPV and NPV) were 82% and 95%, respectively. The classification performance based on AI LVEF < 40% is shown in **Figure 3, c & d**. Site-specific analyses showed consistently high discriminative ability, with AUC values ranging from 0.929 to 0.989. PPV varied between 70% and 97%, while NPV ranged from 94% to 97% across sites. Additional metrics are shown in **Supplementary Table 3**.

**Figure 3.**
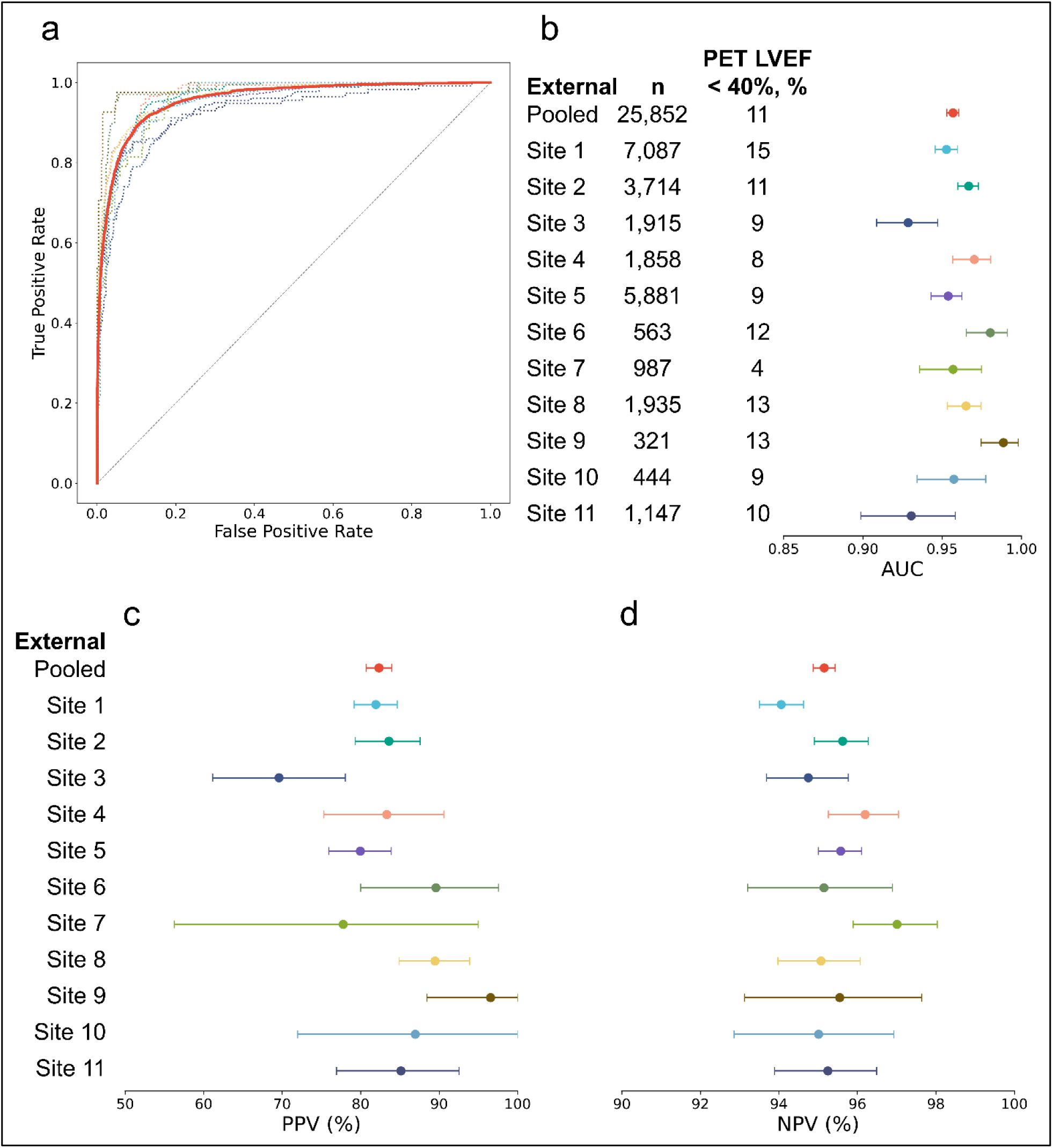
External classification performance for AI CT ejection fraction < 40% Discriminative power of left ventricular ejection fraction (LVEF) derived from ungated chest CT was assessed by the area under the operating characteristic curves (AUCs) in the pooled (red solid line) and site-level (dotted lines) external validation (a). The prevalences of reduced LVEF according to the reference PET LVEF were presented with corresponding AUCs (b). The positive and negative predictive values (PPV, c and NPV, d) were shown for the pooled and site-level external validation.

The AUC for AI LVEF < 50% was 0.934 (95% CI: 0.930–0.937), and that for AI LVEF < 35% was 0.961 (95% CI: 0.957–0.966). The discriminative performance for thresholds of 50% and 35% is shown in **Extended Figure 1**. The per-site classification performance based on the threshold of 50% is shown in **Supplementary Table 4**. The overall PPV and NPV were 80% and 92%, respectively. The per-site classification performance based on the threshold of 35% is shown in **Supplementary Table 5**. The overall PPV and NPV were 82% and 96%, respectively.

In cohort 2, AI LVEF from ungated chest CT aligned well with the echocardiographic LVEF. For LVEF < 40%, the AUC was 0.913 (95% CI: 0.830–0.976), with a PPV of 81% (95% CI: 59%–100%), and an NPV of 96% (95% CI: 91%–99%). For LVEF < 50%, the AUC was 0.933 (95% CI: 0.872–0.978), with a PPV of 79% (95% CI: 62%–93%), and an NPV of 94% (95% CI: 89%–98%). For LVEF < 35%, the AUC was 0.961 (95% CI: 0.902–0.996), with a PPV of 75% (95% CI: 40%–100%) and an NPV of 95% (95% CI: 91%–98%).

### Prognostic evaluation of reduced AI ejection fraction from CT

In cohort 1, over 4.3 ± 3.1 years of follow-up, 2,215 (8.6% of 25,783 patients) heart failure events occurred across 11 imaging centers (**Supplementary Table 6**). An LVEF < 40% from ungated CT-derived LVEF was associated with a 5-fold increase in the risk of heart failure-related hospitalization (hazard ratio [HR] 5.11, 95% CI 4.65-5.62, *P* < 0.001), which was similar to the predictive value of gated PET LVEF (HR 5.09, 95% CI 4.66-5.56, *P* < 0.001; **Figure 4, Heart Failure**).

**Figure 4.**
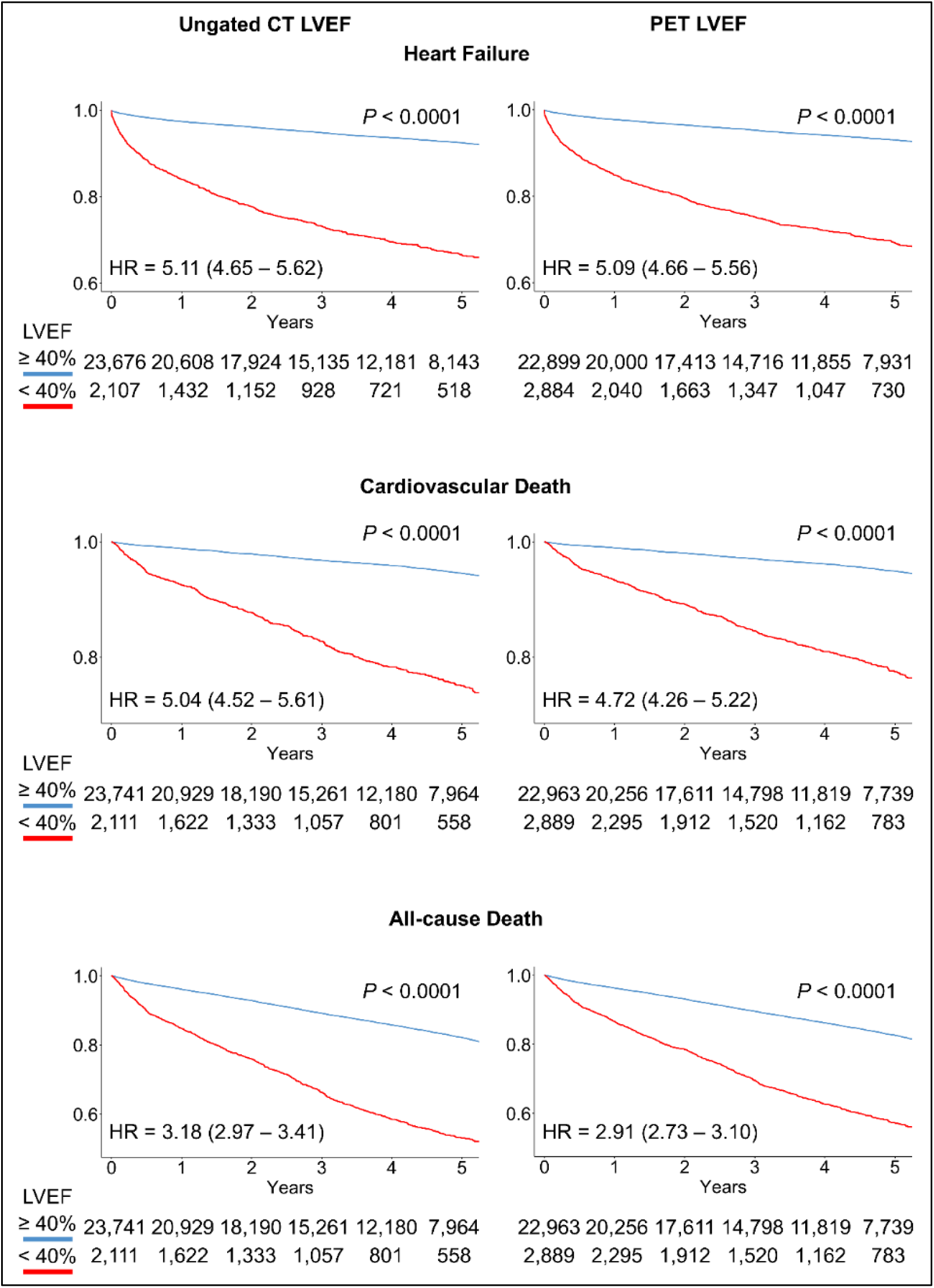
AI LVEF from ungated CT predicts clinical outcomes in cohort 1 Kaplan-Meier curves of left ventricular ejection fraction (LVEF) < 40% (red) against LVEF ≥ 40% (blue) according to the risk of heart failure-related hospitalization, cardiovascular death, and all-cause death in cohort 1 are shown. Unadjusted hazard ratios (HRs) are presented with 95% confidence intervals. P-values from the log-rank test of survival curves are shown above the curves.

In terms of mortality, 5,291 (20%) all-cause death events, including 1,730 (7%) events attributed to cardiovascular disease, occurred for 25,852 patients during the aforementioned period. A < 40% CT-derived LVEF was associated with an over 5-fold increase in the risk of cardiovascular death (HR 5.04, 95% CI 4.52-5.61, *P* < 0.001) and a 3-fold increase for all-cause death (HR 3.18, 95% CI 2.97-3.41, *P* < 0.001). For comparison, the gated PET LVEF had similar risk stratification for cardiovascular death (HR 4.72, 95% CI 4.26-5.22, *P* < 0.001) and all-cause death (HR 2.91, 95% CI 2.73-3.10, *P* < 0.001; **Figure 4, Cardiovascular Death & All-cause Death**).

The association with mortality was further evaluated in the NLST cohort with 24,054 participants (baseline characteristics shown in **Supplementary Table 7**). During 6.4 ± 1.1 years of follow-up, 1,762 (7%) participants died, of which 449 (2%) were categorized as cardiovascular deaths. A < 40% AI LVEF was associated with an over 13-fold increase in the risk of cardiovascular death (HR 13.33, 95% CI 9.67-18.37, *P* < 0.001) and an over 4-fold increase for all-cause death (HR 4.67, 95% CI 3.60-6.05, *P* < 0.001; **Extended Figure 2**).

### Individualized interpretation of model findings

The feature importance analysis from SHapley Additive exPlanations (SHAP)^9^ identified the ungated volumes of the left ventricle (LV) and left ventricular myocardium, heart rate, systolic pressure, and median blood density of the right ventricle (RV) as the highest-ranked features for predicting LVEF (**Extended Figure 3**). After addressing the multicollinearity,^10^ the modified importance order^11^ for LVEF prediction ranked LV volume, LV myocardial volume, heart volume, RV volume, and left atrial volume as the top five features. SHAP enables the derivation of feature importance for individual patients, facilitating personalized interpretation. We present three cases of LVEF prediction in **Figure 5**, featuring CT slices overlapped by CT structures and individually highlighting the top ten features.

**Figure 5.**
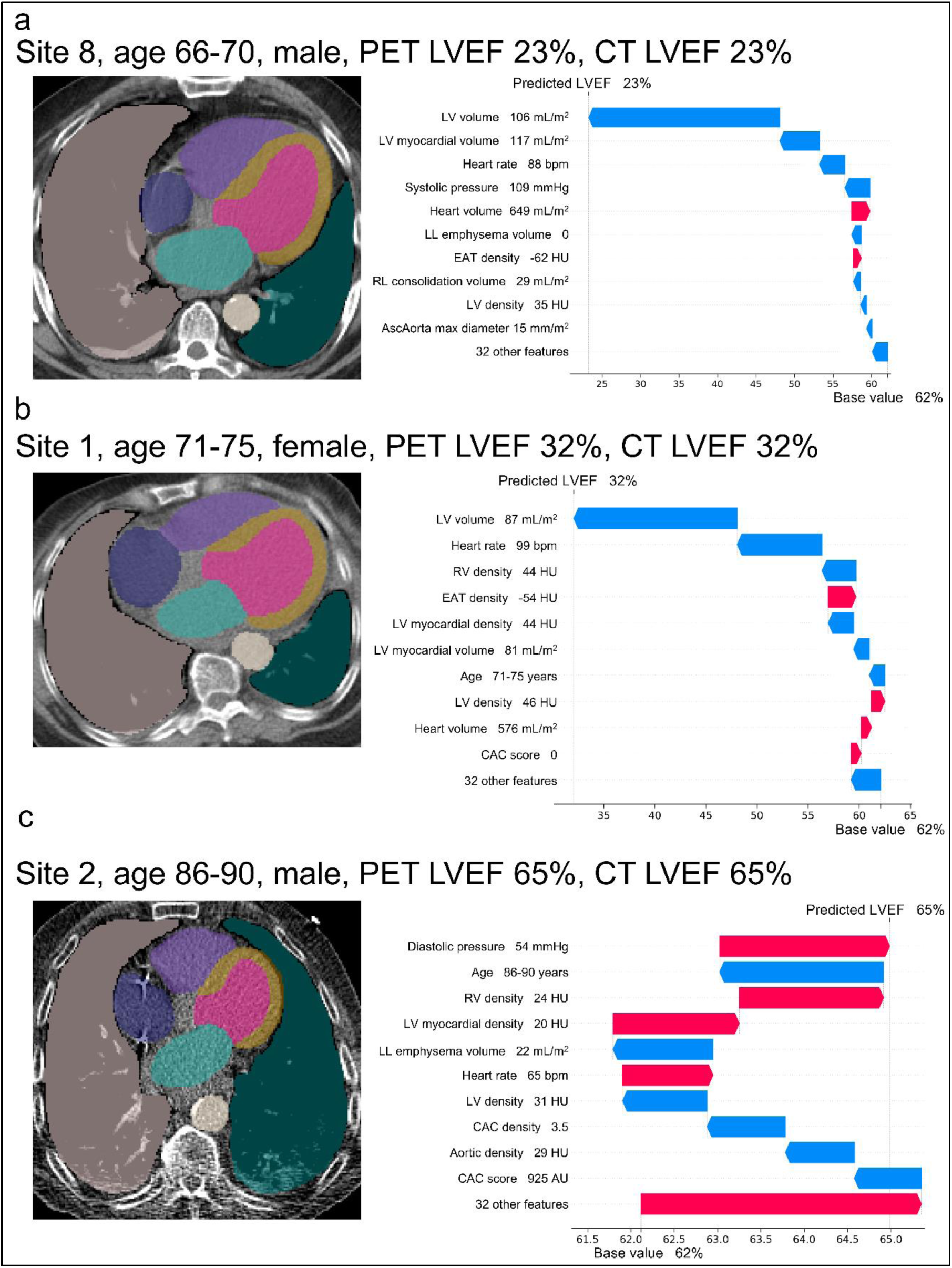
Cases for illustration Case (a) shows a male patient in his 60s from site 8 with an increased left ventricular (LV) volume (106 mL/m2) indexed to body surface area. The patient had a reduced left ventricular ejection fraction (LVEF) of 23% based on PET. The CT prediction was 23%. The factors that increased the potential predicted value (red arrows) included heart volume (649 mL/m^2^), epicardial adipose tissue (EAT) density (-62 Hounsfield Units [HU]). The LVEF prediction was shifted from the base value of 62% to 23%, primarily driven by the LV volume, LV myocardial volume (117 mL/m^2^), heart rate (88 bpm), systolic pressure (109 mmHg), left lung (LL) emphysema volume (0), right lung (RL) consolidation volume (29 mL/m^2^), LV median blood density (35 HU), and ascending aorta (AscAorta) max diameter (15 mm/m^2^) (blue arrows). Case (b) shows a female patient in her 70s from site 1 with an increased LV volume (81 mL/m^2^). The patient had a reduced LVEF of 32% on PET and 32% on our LVEF CT tool. The predicted value was increased by EAT density (-54 HU), LV density (46 HU), heart volume (576 mL/m^2^), and no coronary artery calcium (CAC), and was decreased by LV volume, heart rate (99 bpm), right ventricle (RV) median blood density (44HU), LV myocardial density (44 HU), LV myocardial volume (81 mL/m^2^), and age. Case (c) shows a male patient in his 80s from site 2. The patient had a normal LVEF of 65% based on PET and 65% from prediction. The potential predicted value was decreased by age, left lung emphysema volume (22 mL/m^2^), LV density (31 HU), CAC density (3.5), aortic median blood density (29 HU), and CAC score (925 Agatston Units [AU]), and increased by diastolic pressure (54 mmHg), RV density (24 HU), LV myocardial density (20 HU), and heart rate (65 bpm). Segmented structures shown in chest CT: LV myocardium (yellow), LV (pink), RV (purple), left atrium (turquoise), right atrium (blue), aorta (light grey), left lung (green), right lung (dark grey). Age is presented in ranges to remove patient-identifying information. Actual age values were used in the analysis.

## Discussion

In this study, we demonstrate for the first time that LVEF (traditionally requiring imaging across the cardiac cycle with multiple frames) can be reliably and accurately estimated from ungated, non-contrast, low-dose chest CT scans. We utilize machine-learning algorithm that integrates deep-learning-derived anatomical measurements and clinical variables and can provide individualized explanation. The robustness and generalizability of our approach were confirmed through rigorous external validation across 11 international imaging centers of over 25,000 patients, validating against accurate 3D PET measurments of the LVEF. The results show a strong correlation and high classification accuracy at clinically relevant LVEF thresholds, confirming the clinical utility and reliability across different populations and settings. Furthermore, the AI LVEF accurately stratified patients by risk of heart failure and mortality in this population. Additionally, the prognostic value of the AI LVEF derived from lung CT scans in an independent population of over 24,000 participants from the 33-site lung screening trial showed excellent risk stratification of all cause and cardiovascular mortality.

The predicted LVEF showed a strong correlation with that measured by gated PET, and high AUC, PPV, and NPV for predicting reduced ejection fraction (< 40%) across individual sites and in the pooled analysis, respectively. We found similar results with reduced LVEF thresholds of <35% and <50%. These results indicate strong predictive accuracy and excellent discriminative performance across multiple LVEF thresholds for identifying patients with reduced LVEF. Our comparisons of CT and PET LVEF, and CT and echocardiographic LVEF, align with the intermodality variability in LVEF reported in previous studies. According to a recent meta-analysis^12^, a strong correlation between PET- and cardiac magnetic resonance (CMR)-derived LVEF was observed with a mean difference of less than 1%. Another prospective study reached a similar conclusion, with minimal bias and a limit of agreement of -8% to 7%.^13^ Equivalent findings have been reported for CMR and 2D- or 3D-transthoracic echocardiography (TTE) on 50 patients.^14^ However, the recent prospective MATCH study of over 2100 participants with a median between-scan interval of 1.2 months reported that CMR quantifies cardiac function more reliably than TTE.^15^ Importantly, LVEF values measured by these two imaging methods were significantly different (CMR = 69%, IQI 64%, 74%; 2D-TTE = 58%, IQI 56%, 62%; bias = 11%; Pearson r = 0.40), showing a limited transferability between CMR and TTE. Similar discrepancies have been reported by Pellikka et al in a multicenter randomized trial with over 2000 patients with LVEF ≤ 35% or coronary artery disease (CAD).^16^ The MAE of LVEF between 2D-TTE and CMR was 7.3% with the limit of agreement range exceeding 35%. In view of the aforementioned differences in LVEF measurements between modalities, including gold standards^17^ (CMR and echocardiography), and the extensive validation of the predictions, our proposed ungated non-contrast CT-derived LVEF approach can be positioned as a novel, robust tool for evaluating cardiac function.

Our international cohort of patients undergoing cardiac PET scans presented varying prevalence of reduced LVEF. Derived from the gated PET, the cohort had a median LVEF of 64% (IQI 54%, 72%) with 11% having an LVEF < 40%. While the median of LVEF was close to that of the normal population, the calculated prevalence of reduced LVEF (LVEF < 40%) ranged from 0.01% to 0.11% in the normal population.^18,19^ In the prospective MESA Early HF study, the median LVEF was 63% (IQI 59%, 66%), with 2% having LVEF < 50%.^20^ However, in the test population, the prevalence of reduced LVEF is much higher. In a prospective validation study, 5% of electrocardiography (ECG) patients had an LVEF of ≤35% as determined by TTE.^21^ In another LVEF quantification study, two prospective cohorts were included for external validation, with 10% and 13% having an LVEF of ≤ 40%, respectively.^22^ In these two cohorts, the MAE ranged from 5% to 8%, and the AUC ranged from 0.85 to 0.95. In our leave-one-site-out external validations, the prevalence of LVEF < 40% was lower than 10% in 5 centers (4% to 9%, MAE 5% to 6%, r 0.77 to 0.85, AUC between 0.93 to 0.97, PPV 70% to 83%, NPV 95% to 97%) and equal to or higher in 6 centers (10% to 15%, MAE 5% to 6%, r 0.77 to 0.88, AUC 0.93 to 0.99, PPV 82% to 97%, NPV 94% to 96%). The strong correlation and high classification accuracy of our approach were confirmed in patients with diverse LVEF profiles and different prevalence of reduced LVEF, demonstrating its robustness and generalizability.

CT is the most widely used advanced imaging modality globally, with ungated chest CT performed in approximately 20 million patients in the USA alone.^5^ While chest CT was historically primarily used to diagnose lung abnormalities, it is increasingly recognized that these non-cardiac CT acquisitions can provide important cardiovascular insights. Coronary and large-vessel calcifications are a hallmark of atherosclerosis and, as demonstrated in multiple studies, can be efficiently quantified using non-gated low-dose CT data for risk stratification.^23,24^ Our study further expands the cardiovascular utility of low-dose ungated CT by providing LVEF measurements. Our approach has the advantage of evaluating continuous LVEF, providing an opportunity for a more nuanced diagnosis. Indeed, the diagnosis of HF and the following therapy may depend on varying LVEF thresholds in a sex-specific population.^25,26^ It is also important that patients with recovered LVEF be evaluated at follow-up visits.^27^

Although evaluating LVEF from static chest CT scans without ECG gating may seem counterintuitive, the wealth of data from a chest CT scan allows the derivation of several key biomarkers that, when harnessed with AI, can enable LVEF evaluation. Indeed, by jointly considering chamber and myocardial volumes, the epicardial adipose tissue, coronary calcium, and lung, which can all be fully automatically evaluated from chest CT using deep learning, our approach accurately estimates LVEF.

Contributions of specific biomarkers are displayed in the interpretability analysis using SHAP values and individual patient examples. In our study, chamber volumes, densities of chambers and myocardium, baseline heart rate and blood pressure, coronary calcium, lung, aorta, as well as epicardial adipose tissue, had the largest impact on LVEF. LV volume was the most influential feature in predicting LVEF. Larger LV volumes were consistently associated with lower predicted LVEF, reflecting the established relationship between LV dilation and systolic dysfunction. This finding aligns with the pathophysiology of cardiomegaly, where progressive ventricular remodeling and dilation impair contractile efficiency, leading to reduced LVEF.^2,28^ In contrast, smaller LV volumes were associated with higher LVEF, consistent with preserved or hyperdynamic function. Interestingly, the direction of effect for LV and RV volumes differed: while LV dilation was associated with lower LVEF, smaller RV volumes predicted lower LVEF. This may reflect distinct structural adaptations in left-versus right-sided heart failure phenotypes. In advanced left-sided failure, LV dilation can coexist with RV underfilling and reduced RV preload due to impaired pulmonary circulation^29^, whereas RV dilation often indicates right-sided pressure overload or pulmonary hypertension rather than isolated LV systolic dysfunction.^30^ Besides volume, higher densities of the myocardium and RV were associated with lower LVEF as well, which may be explained by the fibrosis or remodeling of these structures. Heart rate and blood pressure also contributed significantly to the LVEF evaluation. Higher heart rate and lower systolic pressure may reflect the depressed contractility that reduces the forward stroke volume and arterial pressure and increases sympathetic tone.^1^ Diastolic pressure may be elevated due to vasoconstriction, arterial remodeling volume overload.

The predicted LVEF was associated with coronary calcium, epicardial fat, lung, aorta, and pulmonary artery size. Lower CAC scores were linked to higher predicted LVEF, whereas higher CAC scores indicated a greater likelihood of reduced LVEF. This likely reflects the role of coronary atherosclerosis in impairing myocardial perfusion and promoting ischemic cardiomyopathy^31^ or leading to infarction^32^. High CAC scores are associated with greater coronary plaque burden, which is linked to chronic myocardial injury and fibrosis, ultimately leading to ventricular dysfunction.^33,34^ Higher EAT density, while associated with higher mortality risk,^8^ positively contributed to LVEF prediction. Attenuation-based lung volumes were major features as well, particularly volumes of > - 100 HU (for consolidation and denser tissues)^35^ and -700 to -100 HU (for ground-glass opacities)^36^. Together, these features highlight the multifactorial nature of systolic dysfunction, in which structural remodeling, vascular disease burden, hemodynamics, and metabolic alterations contribute to LVEF impairment.

Importantly, in the present study, we reported an explainable machine-learning approach for predicting LVEF from ungated, non-contrast chest CT scans. To our knowledge, this is the first study to automatically quantify LVEF from chest CT. Unlike other studies focusing on modalities already standard for LVEF determination^22^, our approach unlocked the unprecedented potential of ungated, non-contrast chest CT scans for this cornerstone measurement. In our study, CT scans were acquired contemporaneously (within a few minutes) of the gated PET scans, during which LVEF was quantified. Compared to other studies in which the target imaging modality and the gold standard of LVEF were obtained separately with intervals ranging from 1 to 2 months,^37,38^ our predictions were trained and evaluated with the coincident data, minimizing potential differences between scans, thereby achieving a higher accuracy for real-life implementation. We leveraged the multicenter registry to externally evaluate performance across all 11 international imaging centers (total n = 25,852) using the leave-one-site-out strategy. While data drift across patient populations and imaging scanners may hamper performance, the excellent discriminatory and classification performance (AUC 0.96 for LVEF < 40%) demonstrated superior robustness and generalizability, outperforming other studies with fewer external sites and smaller testing cohorts^22^. More importantly, we developed an explainable approach by assessing feature importance to promote interpretability and provide insights into imaging and clinical measurements. The left ventricular size contributed most to LVEF prediction, consistent with clinical evidence.^2^ Lastly, we showed that the predicted LVEF had similar prognostic value compared to PET LVEF, with significant risk stratification of heart failure and mortality.

The data were collected retrospectively from patients undergoing PET MPI at multiple institutions. The inclusion of patients with suspected or established CAD confounded the prevalence of reduced LVEF and the incidence of HF hospitalization. However, we obtained good to excellent agreement for LVEF quantification and classification in external validations with varying prevalence, comparable to other prospective LVEF studies, and demonstrated that CT-derived LVEF had prognostic value similar to LVEF from cardiac PET. We used LVEF from gated cardiac PET as the reference label and evaluated the agreement with echocardiographic LVEF. PET LVEF quantification does not require geometric assumptions, but it may underestimate LVEF due to the low temporal and spatial resolution.^2^ Additional validation with CMR may provide a more accurate evaluation of the performance. The retrospective design of the multicenter registry limits the applicability of this approach in real-world settings. Despite that, we evaluated the agreement in a separate cohort and evaluated the prognostic value of CT LVEF in the prospective NLST dataset. Future prospective studies are desirable.

LVEF can be accurately quantified from non-contrast, ungated, low-dose chest CT scans using an explainable AI approach. The validity was confirmed through external validation across 11 international sites. The CT-derived LVEF measurement proved to be a major prognostic tool, effectively stratifying patients for heart failure and mortality over a 5-year period. This study offers an efficient means of assessing cardiac function with potential widespread clinical ramifications.

## Methods

### Data collection

#### This study utilized 3 separate cohorts

Cohort 1 included patients with cardiac PET/CT from 11 sites in a multicenter, international registry^39^. Consecutive PET/CT scans from each site for patients who underwent PET for evaluation of suspected or known coronary artery disease (CAD) were included to reflect real-world clinical practice. The registry included scans performed between 2007 and 2024. Clinical variables collected at each site were de-identified and anonymized prior to transfer to the core site using dedicated software compliant with the Health Insurance Portability and Accountability Act (HIPAA). The investigators ensured that the institutional ethics committee at each center evaluated and approved the study protocol before data collection and transfer. The overall registry was approved by the Institutional Review Board (IRB) at Cedars-Sinai Medical Center (Office of Research Compliance and Quality Improvement).

Cohort 2 comprised consecutive patients from a single hospital between 2019 and 2024. The inclusion criterion was any patient who underwent non-contrast, ungated chest CT imaging and echocardiography within 180 days. Patients without echocardiography or quantitative LVEF assessment in the echocardiography report were excluded. The Bioethical Committee of the University of Zielona Góra granted a waiver of informed consent documentation.

Cohort 3 included participants from the National Lung Screening Trial (NLST)^40^. The NLST, a multicenter randomized controlled trial, included current or former heavy smokers aged 55 to 74 who were randomly assigned to undergo non-contrast, ungated chest CT imaging between 2002 and 2007. We studied the CT scans of 24,054 participants with available follow-up for death and cause of death. All participants provided written informed consent for participation in the NLST, which was approved by the IRB at each clinical center. The study used de-identified image sets and did not collect new data; therefore, it is considered non-human subject research.

### Ejection fraction quantification from cardiac PET

In cohort 1, acquisition and processing protocols for cardiac PET have been reported previously.^39^ For the gold standard reference and training, LVEF was automatically quantified from 3D electrocardiogram-gated PET using the standard clinical quantitative PET package QPET (Cedars-Sinai, Los Angeles)^41,42^. Only resting PET/CT scans were used in this analysis. PET radiotracers are listed in **Supplementary Table 8**.

### Ejection fraction quantification from echocardiography

In cohort 2, 2D echocardiography was performed using a Philips Epiq 7c, Epiq 7G, or iE33 ultrasound system. LVEF was assessed using biplane or single-plane measurement or visual estimation.^16,19^

### CT acquisition

In cohort 1, chest CT scans were obtained for CT attenuation correction at the same time as the PET scan, with imaging parameters listed in **Supplementary Table 9**. All scans were non-electrocardiographically (ECG) gated and non-contrast enhanced. In cohort 2, non-contrast ungated CT scans were acquired with 2 CT systems from Canon (99%) and United Imaging (1%). All scans were imaged with a peak tube voltage of 120. The reconstruction slice thickness was 0.5 mm (36%), 1mm (63%), or 1.5 mm (1%). In cohort 3, non-contrast ungated chest CT scans were acquired using 17 camera systems, including systems from GE Healthcare (54%), Philips (9%), Siemens (31%), and Toshiba (6%), from 33 screening centers across the United States. Most participants were imaged at a peak tube voltage of 120 (90.2%), followed by 140 (9.75%), and a small number at 100, 130, and 135 (together for < 0.1%). Median tube current was 90 mA (interquartile interval 80–120). Reconstruction thickness ranged from 1–6 mm, with most participants having a slice thickness of 2.5 mm (48%) or 2 mm (32%).

### Segmentation of anatomical structures

We applied established deep learning segmentation models to CT scans to derive measurements of key anatomical structures visible on chest CTs.

#### Segmentation of coronary artery calcium

We used our previously validated model for coronary artery calcium (CAC) segmentation.^43,44^ The segmentation model consists of two networks for the heart silhouette and CAC lesions, respectively, which were tuned for varying scan slice thickness. The heart mask was used for territory correction of CAC candidates in non-cardiac regions. The Agatston score^45^ and density^46^ of CAC were computed. Body-surface area (BSA)^47^-indexed heart volume was computed from the heart mask.

#### Segmentation of cardiac chambers, myocardium, aorta, pulmonary artery, and lung

A pre-trained algorithm, TotalSegmentator^6^, an nn-Unet^48^ based model, was used to segment cardiac chambers (left and right atria, left and right ventricles), left ventricular myocardium, aorta, pulmonary artery, and left and right lungs. Maximal and median diameters of the ascending and descending aorta were extracted.^49,50^ Attenuation-based lung volumes were measured (< -950 HU for emphysema^51,52^, -950 to -700 HU for normal tissue, -700 to -100 HU for ground-glass opacities^36,53^, and > -100 HU for consolidation and denser tissues^35^). BSA-index ungated volumes^54^ and densities (median attenuations) of chambers, left ventricular myocardium, aorta, pulmonary artery, and left and right lungs, and BSA-indexed aortic diameters were computed.

#### Segmentation of epicardial adipose tissue

We used another previously validated model for epicardial adipose tissue (EAT) segmentation. The EAT model was modified from the CAC model by expanding the heart silhouette to the pericardium.^8^ A thresholding of CT attenuation was applied to segment EAT within the heart silhouette.^55^ BSA-indexed volume and density (median attenuation) of EAT were computed.

### Predicting ejection fraction

A boosting regressor (eXtreme Gradient Boosting)^56^ was designed to integrate: measurements of cardiac chambers and myocardium, measurements of aorta, pulmonary artery, and left and right lungs; score and density of CAC; measurements of EAT; heart rate, systolic and diastolic blood pressure; age, sex, height, and weight. The regressor predicts the actual LVEF values. To avoid non-physiological values, the input PET LVEF values above 75% and below 10%. were clipped ^12^. The model for cohort 3 did not include heart rate or blood pressure since these were not available from the NLST dataset.

### Leave-one-site-out external validation

To evaluate the robustness of the approach in external settings, external validation was performed using a repeated leave-one-site-out (LOSO) framework across 11 participating sites of cohort 1. In this approach, the models were trained exclusively on data from 10 sites and tested on the remaining site. This process was repeated, yielding 11 separate models, with each model independently tested in a separate external site. Performance metrics from these evaluations were then concatenated to yield pooled estimates of performance, ensuring generalizability across heterogeneous clinical settings. This approach avoids random selection of training and testing sites that could lead to biased results.^57^

### Evaluation of continuous LVEF prediction

The regression performance of LVEF prediction was evaluated using multiple complementary metrics. Bias was calculated as the difference between the true LVEF values and the predicted LVEF values. Mean absolute error (MAE) was used to quantify the average magnitude of prediction error. Agreement between predicted and reference LVEF was further assessed using the intraclass correlation coefficient (ICC), Pearson’s correlation coefficient (r), and coefficient of variation. Values were presented with 95% confidence intervals (CIs).

### Clinical thresholds and evaluation of binary classification

For evaluation of clinical utility, performance in the prediction of reduced LVEF (< 40%)^1^ was evaluated using the area under the receiver operating characteristic curve (AUC), positive predictive value (PPV), negative predictive value (NPV), sensitivity, and specificity. As secondary endpoints, we also evaluated the prediction of LVEF < 50% and 35%.^1^ Values were presented with 95% CIs.

### Agreement with echocardiographic LVEF

To evaluate the agreement with LVEF derived from echocardiogram, continuous and reduced (< 40%) AI LVEF from a model trained with the entire cohort 1 were compared with echocardiographic LVEF in cohort 2. Values were presented with 95% CIs.

### Feature importance

We applied SHapley Additive exPlanations (SHAP)^9^ to the regressor trained with the entire cohort 1 to rigorously assess feature importance and ensure interpretability. SHAP provides both local (individual prediction) and global (overall model behavior) insights into how imaging and clinical variables influence predictions, supporting transparent clinical decision-making. We leveraged both local and global explainability to delineate personalized risk interpretations and population risk profiles. To address the collinearity within features, we applied the Modified Index Position (MIP) to modify the ranking of features with strong correlations that were assigned low SHAP scores despite being significantly associated with LVEF.^10,11^ We also evaluated the stability of the feature ranks using the Normalized Movement Rate (NMR).^58^

### Prognostic validation

We also performed the prognostic evaluation in cohort 1 and cohort 3 for the ungated CT LVEF. In cohort 1, patients were followed for HF based on hospitalization and death. Hospitalization for HF was defined by the primary discharge diagnosis, determined through previously validated international classification of diseases (ICD) codes. Follow-up for HF was performed locally at each imaging center. HF was obtained from hospital electronic medical records (centers in the United States and Europe) or patient contact (centers in the United States). Death was retrieved from the Social Security Death Index (centers in the United States), from chart review at the site or from patient contact (Mexico), and from the hospital electronic medical records (Europe). Cardiovascular deaths included those attributed to myocardial infarction, sudden cardiac death, heart failure, stroke, cardiovascular procedures, cardiovascular hemorrhage, or other cardiovascular causes.^59^ Cardiovascular death was determined by review of the death certificate, hospital chart, or physician’s records. In cohort 3, participants were followed for death, and information from death certificates regarding the underlying cause. Cardiovascular death was determined for the ICD-10 codes using established definitions for cardiovascular death.^59^ A < 40% threshold for LVEF derived from gated PET or the CT prediction (pooled across sites for cohort 1 and from a model trained with the entire cohort 1 for cohort 3) was used to separate patients into two risk groups. Cumulative event rates of HF, cardiovascular death, and death were computed with the Kaplan-Meier method. Comparisons of survival curves were performed with the log-rank test. We used the unadjusted Cox proportional hazards regression to assess the stratification value of predicted LVEF. Hazard ratios (HRs) were presented with 95% CIs.

### Statistics

Continuous variables are presented as mean ± standard deviation for normally distributed data and median (interquartile interval) for non-normally distributed data. A two-tailed p-value < 0.05 was considered statistically significant. Statistical analyses were performed using R (Version 4.4.2, R Foundation for Statistical Computing, Vienna, Austria) and Python 3.10.13.

## Data availability

Original data from the NLST can be requested through the National Cancer Institute for specific projects. To the extent allowed by data sharing agreements and IRB protocols, data from this manuscript will be shared upon written request.

## Author contributions

J.Z., D.D. and P.J.S. designed the study. K.P., G.R., M.L., R.J.H.M., P.B.K., J.X.L., L.S., M.I.T., E.A., I.C-J., R.R.S.P., M.A.-M., A.J.E., W.A., S.K., V.T.L., S.M., T.L.R., J.H., S.W., P.C., D.S.B., M.F.D.C. and P.J.S. collected and organized data. J.Z., A.S., G.R., M.L., A.K., R.J.H.M., P.B.K. and J.X.L. curated the dataset. P.J.S. and J.Z. developed the AI prediction code. J.Z. performed statistical tests and created all figures. J.Z., J.K. and P.J.S. drafted the manuscript with feedback from all authors. P.J.S. supervised the study. All authors discussed the results and approved the final version before submission.

## Acknowledgements

This research was supported by grants R01EB034586 and R35HL161195 from the National Institute of Biomedical Imaging and Bioengineering and the National Heart, Lung, and Blood Institute (Principal Investigator, Piotr Slomka). The authors thank the National Cancer Institute for access to National Cancer Institute (NCI) data collected by the National Lung Screening Trial (NLST) under project number NLST-981. The content is solely the responsibility of the authors and does not necessarily represent the official views of the National Institutes of Health.

## Extended Data

**Extended Figure 1.**
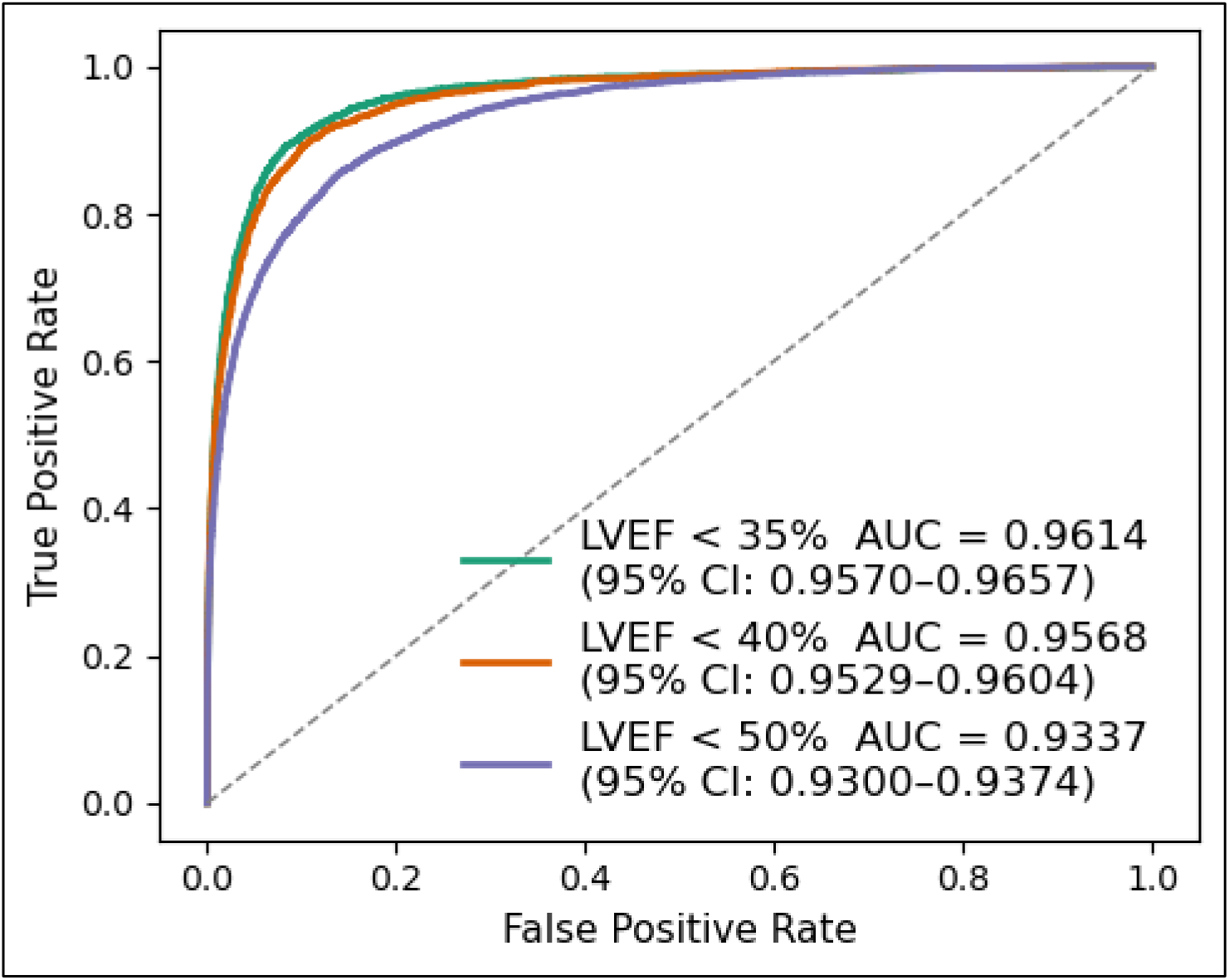
Discrimination performance for different LVEF thresholds Discriminative power of left ventricular ejection fraction (LVEF) derived from ungated chest CT was assessed using the area under the operating characteristic curves (AUCs) in the pooled external validation. AUCs were presented with 95% confidence intervals (CIs).

**Extended Figure 2.**
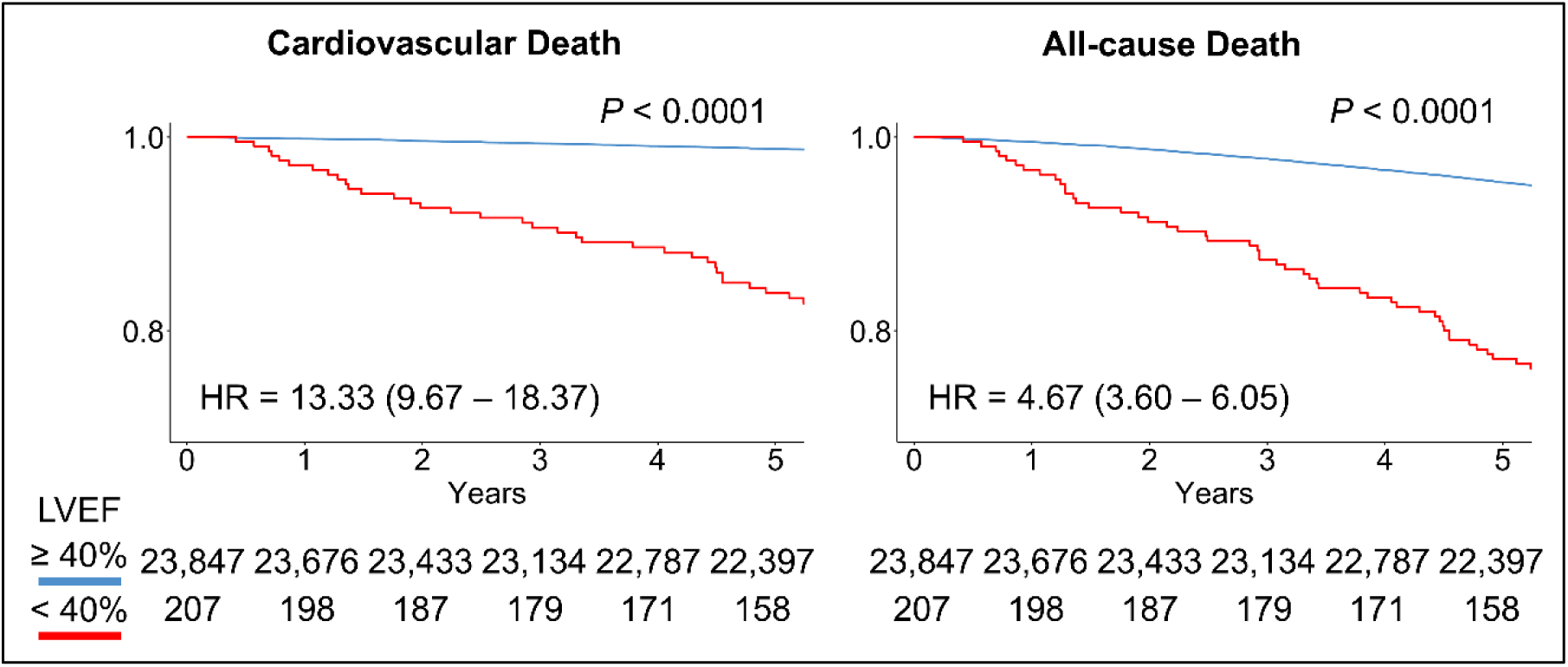
AI LVEF from ungated CT predicts clinical outcomes in cohort 3 Kaplan-Meier curves of left ventricular ejection fraction (LVEF) < 40% (red) against LVEF ≥ 40% (blue) according to the risk of cardiovascular death (a) and all-cause death (b) in cohort 3 are shown. Unadjusted hazard ratios (HRs) are presented with 95% confidence intervals. P-values from the log-rank test of survival curves are shown above the curves.

**Extended Figure 3.**
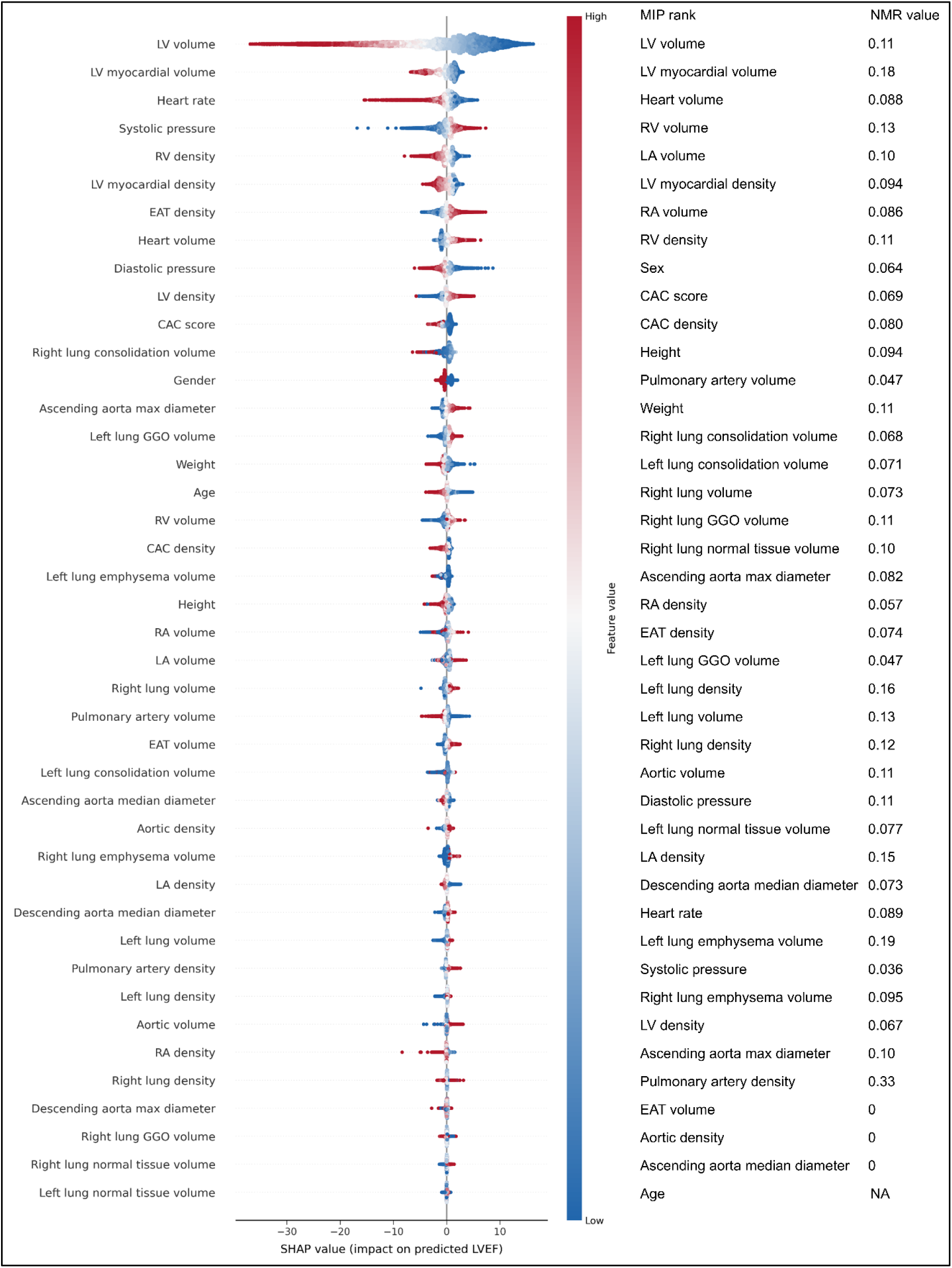
Feature ranking and influence Features used to predict left ventricular ejection fraction (LVEF) are shown in the order of mean absolute SHAP values. Each dot on each feature row corresponds to an instance that provides the matched explanation, with its color indicating the feature value (blue – low, red – high) and its x position indicating the impact on LVEF prediction (positive – increase, negative – decrease). Larger LV (higher values of volume, represented in red color) showed negative impacts on the prediction, which reduced the derived LVEF. The systolic pressure was another important feature. Lower systolic pressure (represented in blue color) was associated with a lower predicted LVEF. Lower predicted LVEF values were also associated with several other factors, including enlarged LV myocardium, raised heart rate, higher median blood density of the right ventricle (RV) and LV myocardium, lower density of EAT and LV, smaller heart size, higher diastolic pressure, and a higher CAC score. To address multicollinearity, the Modified Index Position (MIP) method iteratively removes the top-performing feature and recalculates SHAP values in a new model with fewer features. Normalized Movement Rate (NMR) values are shown for the stability of the feature ranking after removing the corresponding feature. CAC, coronary artery calcium; EAT, epicardial adipose tissue; GGO, ground-glass opacity; LA, left atrium; RA, right atrium.

