## Supplementary Materials for "Ejection fraction quantification from ungated chest CT by AI"

**Supplementary Information**

Supplementary Table 1. Baseline characteristics of cohort 2

| **Characteristic** | **Overall (n = 128)** |
| --- | --- |
| Age (year, mean ± SD) | 67 ± 10 |
| Female, n (%) | 56 (44%) |
| Hemodynamic measurements | |
| Heart rate, beats per minute | 74 (64, 78) |
| Systolic pressure, mmHg | 130 (120, 140) |
| Diastolic pressure, mmHg | 80 (70, 85) |
| Echocardiogram measurement | |
| Ejection fraction (%) | 55 (50, 60) |
| Ejection fraction ≥ 40%, n (%) | 110 (86%) |
| Ejection fraction < 40%, n (%) | 18 (14%) |

Continuous variables are presented as mean ± standard deviation (SD) or median (interquartile interval). Categorical variables are presented as counts (percentages).

Supplementary Table 2. Distribution of patients by PET ejection fraction

| Site | n | LVEF < 50%, n(%) | LVEF < 40%, n(%) | LVEF < 35%, n(%) |
|---|---|---|---|---|
| All | 25,852 | 5,205 (20%) | 2,889 (11%) | 2,034 (8%) |
| Site 1 | 7,087 | 1,805 (25%) | 1,035 (15%) | 754 (11%) |
| Site 2 | 3,714 | 729 (20%) | 409(11%) | 297 (8%) |
| Site 3 | 1,915 | 379 (20%) | 181 (9%) | 123 (6%) |
| Site 4 | 1,858 | 257 (14%) | 147 (8%) | 102 (5%) |
| Site 5 | 5,881 | 982 (17%) | 554 (9%) | 393 (7%) |
| Site 6 | 563 | 117 (21%) | 68 (12%) | 39 (7%) |
| Site 7 | 987 | 121 (12%) | 43 (4%) | 14 (1%) |
| Site 8 | 1,935 | 437 (23%) | 256 (13%) | 182 (9%) |
| Site 9 | 321 | 66 (21%) | 41 (13%) | 30 (9%) |
| Site 10 | 444 | 81 (18%) | 41 (9%) | 25 (6%) |
| Site 11 | 1,147 | 231 (20%) | 114 (10%) | 75 (7%) |

Supplementary Table 3. Classification performance metrics for ejection fraction < 40%

| External validation | AUC | PPV (%) | NPV (%) | Sensitivity (%) | Specificity (%) |
|---|---|---|---|---|---|
| Pooled | 0.957 (0.953–0.960) | 82.3 (80.7–84.0) | 95.2 (94.9–95.4) | 60.2 (58.4–61.9) | 98.4 (98.2–98.5) |
| Site 1 | 0.953 (0.946–0.960) | 81.9 (79.2–84.7) | 94.1 (93.5–94.6) | 64.0 (61.1–66.8) | 97.6 (97.2–98.0) |
| Site 2 | 0.967 (0.960–0.973) | 83.6 (79.3–87.6) | 95.6 (94.9–96.3) | 63.6 (58.7–68.1) | 98.5 (98.0–98.9) |
| Site 3 | 0.929 (0.909–0.947) | 69.6 (61.2–78.0) | 94.7 (93.7–95.8) | 48.1 (40.4–55.8) | 97.8 (97.1–98.5) |
| Site 4 | 0.970 (0.957–0.981) | 83.3 (75.3–90.6) | 96.2 (95.3–97.0) | 54.4 (46.6–62.3) | 99.1 (98.6–99.5) |
| Site 5 | 0.954 (0.943–0.962) | 79.9 (75.9–83.9) | 95.6 (95.0–96.1) | 56.1 (51.9–60.2) | 98.5 (98.2–98.8) |
| Site 6 | 0.980 (0.965–0.991) | 89.6 (80.0–97.6) | 95.1 (93.2–96.9) | 63.2 (50.8–74.6) | 99.0 (98.0–99.8) |
| Site 7 | 0.957 (0.936–0.975) | 77.8 (56.2–95.0) | 97.0 (95.9–98.0) | 32.6 (19.1–47.1) | 99.6 (99.1–99.9) |
| Site 8 | 0.965 (0.953–0.975) | 89.5 (84.9–93.9) | 95.1 (94.0–96.1) | 66.4 (60.7–72.3) | 98.8 (98.3–99.3) |
| Site 9 | 0.989 (0.975–0.998) | 96.6 (88.5–100.0) | 95.5 (93.1–97.6) | 68.3 (53.2–82.5) | 99.6 (98.9–100.0) |
| Site 10 | 0.957 (0.934–0.978) | 87.0 (72.0–100.0) | 95.0 (92.9–96.9) | 48.8 (34.2–64.3) | 99.3 (98.3–100.0) |
| Site 11 | 0.930 (0.899–0.958) | 85.1 (76.9–92.5) | 95.2 (93.9–96.5) | 55.3 (46.4–64.4) | 98.9 (98.3–99.5) |

The discriminative power of ejection fraction derived from ungated chest CT was assessed at 40% in pooled and site-level external validations using the area under the receiver operating characteristic curve (AUC), positive predictive value (PPV), negative predictive value (NPV), sensitivity, and specificity. Values are shown with 95% confidence intervals.

Supplementary Table 4. Classification performance metrics for ejection fraction < 50%

| External validation | AUC | PPV (%) | NPV (%) | Sensitivity (%) | Specificity (%) |
|---|---|---|---|---|---|
| Pooled | 0.934 (0.930–0.937) | 80.0 (78.8–81.2) | 92.0 (91.6–92.4) | 67.0 (65.7–68.2) | 95.8 (95.5–96.1) |
| Site 1 | 0.931 (0.924–0.937) | 81.2 (79.3–83.2) | 90.6 (89.8–91.4) | 71.2 (69.2–73.3) | 94.4 (93.8–95.0) |
| Site 2 | 0.938 (0.929–0.947) | 78.4 (75.3–81.6) | 92.5 (91.5–93.4) | 68.3 (65.0–71.7) | 95.4 (94.7–96.1) |
| Site 3 | 0.903 (0.886–0.920) | 72.6 (67.6–77.6) | 90.7 (89.3–92.1) | 60.9 (56.1–65.7) | 94.3 (93.2–95.5) |
| Site 4 | 0.955 (0.942–0.966) | 78.6 (72.9–84.0) | 94.6 (93.6–95.7) | 65.8 (59.9–71.5) | 97.1 (96.3–97.9) |
| Site 5 | 0.937 (0.929–0.945) | 79.5 (76.6–82.3) | 93.0 (92.2–93.6) | 63.4 (60.3–66.4) | 96.7 (96.2–97.2) |
| Site 6 | 0.946 (0.922–0.968) | 84.2 (77.0–90.8) | 93.1 (90.6–95.2) | 72.6 (64.3–80.6) | 96.4 (94.7–98.0) |
| Site 7 | 0.906 (0.877–0.931) | 74.3 (64.0–84.2) | 92.8 (91.0–94.4) | 45.5 (36.8–54.2) | 97.8 (96.8–98.7) |
| Site 8 | 0.946 (0.934–0.956) | 85.7 (82.0–89.3) | 91.6 (90.2–93.0) | 69.8 (65.2–74.1) | 96.6 (95.6–97.5) |
| Site 9 | 0.961 (0.936–0.982) | 89.3 (80.4–96.7) | 94.0 (90.9–96.6) | 75.8 (64.9–85.7) | 97.6 (95.5–99.2) |
| Site 10 | 0.954 (0.934–0.972) | 80.6 (71.0–89.7) | 92.8 (90.1–95.2) | 66.7 (55.7–76.5) | 96.4 (94.5–98.3) |
| Site 11 | 0.884 (0.858–0.910) | 77.2 (70.7–83.5) | 89.6 (87.7–91.5) | 55.8 (49.6–62.4) | 95.9 (94.5–97.1) |

The discriminative power of ejection fraction derived from ungated chest CT was assessed at 50% in pooled and site-level external validations using the area under the receiver operating characteristic curve (AUC), positive predictive value (PPV), negative predictive value (NPV), sensitivity, and specificity. Values are shown with 95% confidence intervals.

Supplementary Table 5. Classification performance metrics for ejection fraction < 35%

| External validation | AUC | PPV | NPV | Sensitivity | Specificity |
|---|---|---|---|---|---|
| Pooled | 0.961 (0.957–0.966) | 81.7 (79.6–83.7) | 96.3 (96.1–96.6) | 55.7 (53.4–57.8) | 98.9 (98.8–99.1) |
| Site 1 | 0.957 (0.949–0.964) | 82.2 (78.9–85.5) | 95.0 (94.5–95.5) | 56.5 (52.9–60.1) | 98.5 (98.3–98.8) |
| Site 2 | 0.973 (0.967–0.979) | 83.0 (77.9–87.9) | 96.5 (95.9–97.1) | 59.3 (53.7–64.4) | 98.9 (98.6–99.3) |
| Site 3 | 0.938 (0.914–0.958) | 74.1 (62.1–85.2) | 95.7 (94.8–96.6) | 35.0 (26.2–43.6) | 99.2 (98.7–99.6) |
| Site 4 | 0.967 (0.948–0.981) | 80.3 (70.0–90.0) | 97.1 (96.3–97.8) | 48.0 (38.7–57.3) | 99.3 (98.9–99.7) |
| Site 5 | 0.956 (0.943–0.967) | 81.3 (76.8–85.7) | 97.0 (96.5–97.4) | 56.5 (51.4–61.4) | 99.1 (98.8–99.3) |
| Site 6 | 0.983 (0.972–0.991) | 78.1 (62.1–92.0) | 97.4 (95.9–98.7) | 64.1 (48.7–79.3) | 98.7 (97.5–99.6) |
| Site 7 | 0.961 (0.928–0.986) | 41.7 (13.3–71.4) | 99.1 (98.5–99.6) | 35.7 (11.1–63.6) | 99.3 (98.7–99.8) |
| Site 8 | 0.976 (0.967–0.984) | 86.7 (80.7–92.1) | 96.4 (95.5–97.2) | 64.3 (57.2–71.6) | 99.0 (98.5–99.4) |
| Site 9 | 0.976 (0.952–0.992) | 85.0 (68.4–100.0) | 95.7 (93.3–97.7) | 56.7 (39.1–73.9) | 99.0 (97.6–100.0) |
| Site 10 | 0.963 (0.932–0.988) | 82.4 (61.5–100.0) | 97.4 (95.8–98.8) | 56.0 (35.0–76.2) | 99.3 (98.3–100.0) |
| Site 11 | 0.934 (0.896–0.967) | 81.2 (70.0–91.5) | 96.7 (95.7–97.7) | 52.0 (41.0–63.6) | 99.2 (98.6–99.6) |

The discriminative power of ejection fraction derived from ungated chest CT was assessed at 35% in pooled and site-level external validations using the area under the receiver operating characteristic curve (AUC), positive predictive value (PPV), negative predictive value (NPV), sensitivity, and specificity. Values are shown with 95% confidence intervals.

Supplementary Table 6. Number of patients with heart failure

| Site (Number of patients with HF follow-up) | Number of patients with heart failure (%) |
|---|---|
| All (25,783) | 2,215 (8.6%) |
| Site 1 (7,087) | 675 (9.5%) |
| Site 2 (3,645) | 456 (13%) |
| Site 3 (1,915) | 208 (11%) |
| Site 4 (1,858) | 179 (9.6%) |
| Site 5 (5,881) | 313 (5.3%) |
| Site 6 (563) | 26 (4.6%) |
| Site 7 (987) | 44 (4.5%) |
| Site 8 (1,935) | 236 (12%) |
| Site 9 (321) | 36 (11%) |
| Site 10 (444) | 12 (2.7%) |
| Site 11 (1147) | 30 (2.6%) |

Supplementary Table 7. Baseline characteristics of cohort 3

| **Characteristic** | **Overall (n = 24,054)** |
| --- | --- |
| Age (year, mean ± SD) | 61 ± 5 |
| Female, n (%) | 9,772 (41%) |
| Body-mass index ($kg/m^2$, mean ± SD) | 28 ± 5 |
| Smoking (pack years) | 48 (39, 66) |
| Medical history, n (%) | |
| Hypertension | 8,436 (35%) |
| Diabetes | 2,322 (10%) |
| Heart disease | 3,117 (13%) |
| COPD | 1,226 (5%) |
| Stroke | 666 (3%) |

Continuous variables are presented as mean ± standard deviation (SD) or median (interquartile interval). Categorical variables are presented as counts (percentages).

COPD, chronic obstructive pulmonary disease

Supplementary Table 8. Use of PET radiotracers in cohort 1

| **Site** | **Rubidium-82, n (%)** | **Ammonia-13, n (%)** |
|---|---|---|
| All (25,783) | 17,123 (66%) | 8,729 (34%) |
| Site 1 (7,087) | 4,079 (58%) | 3,008 (42%) |
| Site 2 (3,714) | 3,714 (100%) | 0 |
| Site 3 (1,915) | 0 | 1,915 (100% |
| Site 4 (1,858) | 1,858 (100%) | |
| Site 5 (5,881) | 5,881 (100%) | |
| Site 6 (563) | 0 | 563 (100%) |
| Site 7 (987) | 0 | 987 (100%) |
| Site 8 (1,935) | 0 | 1,935 (100%) |
| Site 9 (321) | 0 | 321 (100%) |
| Site 10 (444) | 444 (100%) | 0 |
| Site 11 (1147) | 1,147 (100%) | 0 |

Supplementary Table 9. CT imaging parameters per site in cohort 1.

| Site | Scanner system | Slice thickness, mm | Tube current, mA | Tube voltage, kVp | Breath holding |
|---|---|---|---|---|---|
| Site 1 | GE Discovery MI<br>GE Discovery MI RX<br>GE Discovery MI STE | 2.5-5 | 10-26 | 120-140 | Shallow |
| Site 2 | Siemens Biograph 64 TruePoint<br>Siemens Biograph 128 Vision Edge<br>GE Discovery 710 | 3 | 11-13 | 100 | Shallow |
| Site 3 | Siemens Biograph 64 mCT Flow | 3 | 30 | 120 | Free |
| Site 4 | Siemens Biograph 600 Vision Edge | 3 | 20-50 | 100-120 | Free |
| Site 5 | Siemens Biograph 16 TruePoint<br>Siemens Biograph 20 mCT<br>Siemens Biograph 40 mCT | 2 | 15-38 | 120-130 | Free |
| Site 6 | GE Discovery MI | 3.75 | 14-75 | 120 | Free |

| | | | | | |
|---|---|---|---|---|---|
| Site 7 | Philips Gemini TF TOF 16 | | | | |
| | Philips Gemini TF TOF 64 | 3 | 110-185 | 120 | Free |
| Site 8 | GE Discovery 710 | 3.75 | 17-77 | 120 | Shallow |
| Site 9 | Siemens Biograph 64 TruePoint | | | | |
| | Siemens Biograph Vision 600 | 3 | 33-580 | 120 | Breath hold |
| Site 10 | Philips Ingenuity TF | | | | |
| | GE Discovery Mi | 3 | 35-100 | 120-140 | Free |
| Site 11 | Siemens Biograph 64 mCT | | | | |
| | Siemens Biograph 64 Vision 600 | 3 | 70-200 | 120 | Free |
